# Bowel Colonization by Non-Commensal Fungi in Neonatal Obstructive Cholangitis and Biliary Atresia

**DOI:** 10.1101/2023.03.09.23287026

**Authors:** Song-Wei Huang, Chia-Ray Lin, Ya-Hui Chang, Yen-Hsuan Ni, Huey-Ling Chen, Hong-Hsing Liu

**Author notes:** Equal contribution as first author (Song-Wei Huang, Chia-Ray Lin). Equal contribution as senior author (Huey-Ling Chen, Hong-Hsing Liu).

## Abstract

Up to two-thirds of biliary atresia (BA) patients need liver transplantation after the standard Kasai portoenterostomy treatment. Unrelenting cholangitis often precedes the full-blown presentation of BA. Using ribosomal 18S sequencing, here we report bowel colonization by non-commensal *Aspergillus* fungi in one case of neonatal obstructive cholangitis. Continuous oral fluconazole treatment resolved obstructive cholangitis completely. Colonization by non-commensal *Aspergillus* and *Cerrena* were additionally identified in 2 BA cases. In brief, bowel colonization by non-commensal fungi could be a treatable cause of neonatal obstructive cholangitis and potentially BA at an early stage.

## Introduction

Inflammation of bile ducts or cholangitis in newborns is uncommon. However, it can be a devastating prelude to biliary atresia (BA), the pathologic condition of obliterated bile outlets. BA patients have characteristically low fecal bile pigments at disease onset, and eventually develop clay-colored stools. Kasai portoenterostomy, which surgically rechannels between interhepatic bile ducts and intestine, is currently the standard first-line treatment for BA[1]. However, up to two-thirds of patients will still require liver transplantation[2] due to unrelenting bile-duct obstruction and subsequent liver cirrhosis. The underlying cause for BA has remained unknown.

Here we describe a case of neonatal obstructive cholangitis associated with colonization by non-commensal *Aspergillus* fungi. Oral anti-fungal fluconazole successfully resolved cholangitis and normalized bilirubin levels. Drawing from these findings, we suspected that BA could share a similar pathogenic mechanism. Indeed, feces of two BA patients showed colonization by non-commensal *Cerrena* and *Aspergillus* fungi. Healthy controls showed colonization only by commensal fungi *Candida sp*. and *Malassezia furfur*.

## Methods

### Human subjects

Use of clinical samples and report of clinical data have been approved by the Institutional Review Boards of National Taiwan University Children’s Hospital and En Chu Kong Hospital.

### Fecal DNA extraction

Fecal samples from the index case were collected and stored at −80°C before use. DNA was extracted from 50-200mg stool using ZymoBIOMICS DNA Miniprep Kit (Zymo, D4300) according to manufacturer’s protocols. Fecal DNA for BA cases and controls were extracted with QIAamp PowerFecal Pro DNA Kits.

### 18S PCR and Sanger sequencing

All polymerase chain reactions (PCR) were performed with KAPA HiFi HotStart ReadyMix PCR Kit (Roche, KK2602). For 18S V1-V8 PCR, DNA samples were run with primers 18S-F-bc1005C-P5 (5’-AATGATACGGCGACCACCGAGATCTACACGTGAG CTGAGAGCGCACCATGCATGTCTAAGTWTAA -3’) and 18S-R-bc1033C-P7 (5’-CAAGCAG AAGACGGCATACGAGATTCTCTGACGCTGCTCTAICCATTCAATCGGTAIT -3’). PCR was performed on MiniAmp plus Thermal Cycler (Thermo Fisher Scientific, A37835) with the following conditions: 95°C 3m, 5 cycles of 98°C 30s + 54°C 30s + 72°C 1m30s, 5 cycles of 98°C 30s + 51°C 30s + 72°C 1m30s, 30 cycles of 98°C 30s + 48°C 30s + 72°C 1m30s, and a final extension of 72°C 3m30s. Yeast cDNA and ddH_2_O were used as positive and negative controls, respectively. For semi-nested 18S V7-V8 PCR, the first V1-V8 PCR was performed as above except with the last 30 cycles before final extension reduced to 20 cycles. 1μl of V1-V8 PCR products was used to template each semi-nested V7-V8 PCR with primers 18S-F-nu-SSU-1333-5’ (5’-CGWTAACGAACGAGACCT-3’) and 18S-R-bc1033C-P7. Thermocycling conditions were 95°C 3m, 5 cycles of 98°C 30s + 54°C 30s + 72°C 1m, 5 cycles of 98°C 30s + 51°C 30s + 72°C 1m, 20 cycles of 98°C 30s + 48°C 30s + 72°C 1m, and a final extension of 72°C 3m30s. Semi-nested V7-V8 amplicons were purified with MinElute PCR Purification Kit (QIAGEN, #28006) before Sanger sequencing.

### 16S PCR

All 16S PCR were performed with KAPA HiFi HotStart ReadyMix PCR Kit (Roche, KK2602). V1-V9 regions were amplified with primers 16S-F-bc1005-P5 (5’-AATGATACGGCGACCACCGAGATCTACACCACTCGACTCTCGCGTAGRGTTYGATYMTGG CTCAG-3’) and 16S-R-bc1033-P7 (5’-CAAGCAGAAGACGGCATACGAGATAGAGACTGCGACGAGARGYTACCTTGTTACGACTT-3’) using the following thermocycling conditions: 95 °C 3m, 27 cycles of 95°C 30s + 57°C 30s + 72°C 1m, and a final extension of 72°C 3m. E. coli DH5α DNA and ddH_2_O were used as positive and negative controls, respectively.

### Bioinformatic analyses

Sequences from Sanger sequencing were analyzed with SnapGene software (www.snapgene.com). Nucleotide BLAST[3] was performed using the National Center for Biotechnology Information server.

## Results

### Fungal cholangitis in a newborn

A newborn male was delivered emergently by Caesarean section (C/S) due to fetal distress at 39 weeks of gestation. Cardiac arrest and thick meconium stains were noted. After successful resuscitation, the patient was sent to Neonatal Intensive Care Unit (NICU) for further management. Severe hypoglycemia (< 2 mg/dL) was detected at admission. Continuous supply of 10% glucose via peripheral lines barely maintained blood glucose levels. Without central lines, early oral feeding with 50% concentrated glucose water was begun one day after birth (D1) for one week. Insulin and cortisol responses were normal as evaluated on D3, and there was no evidence of inborn errors of metabolism. Vital signs were stable and no neural sequelae were found. However, ileus with distended abdomen later complicated the course in spite of full antibiotic coverage. Severe thrombocytopenia with normal counts of white blood cells was noted (D8, D11, Figure 1A). Fungal infection was suspected, so intravenous (i.v.) fluconazole was administered from D11. Dramatic clinical improvement followed, and fluconazole was discontinued on D19. However, severe leukocytosis of 20,400/μL on D15 and 28,300/μl on D19 required another empirical course of i.v. antibiotics (D19-D26), but in vain (D27, Figure 1A).

**Figure 1.**
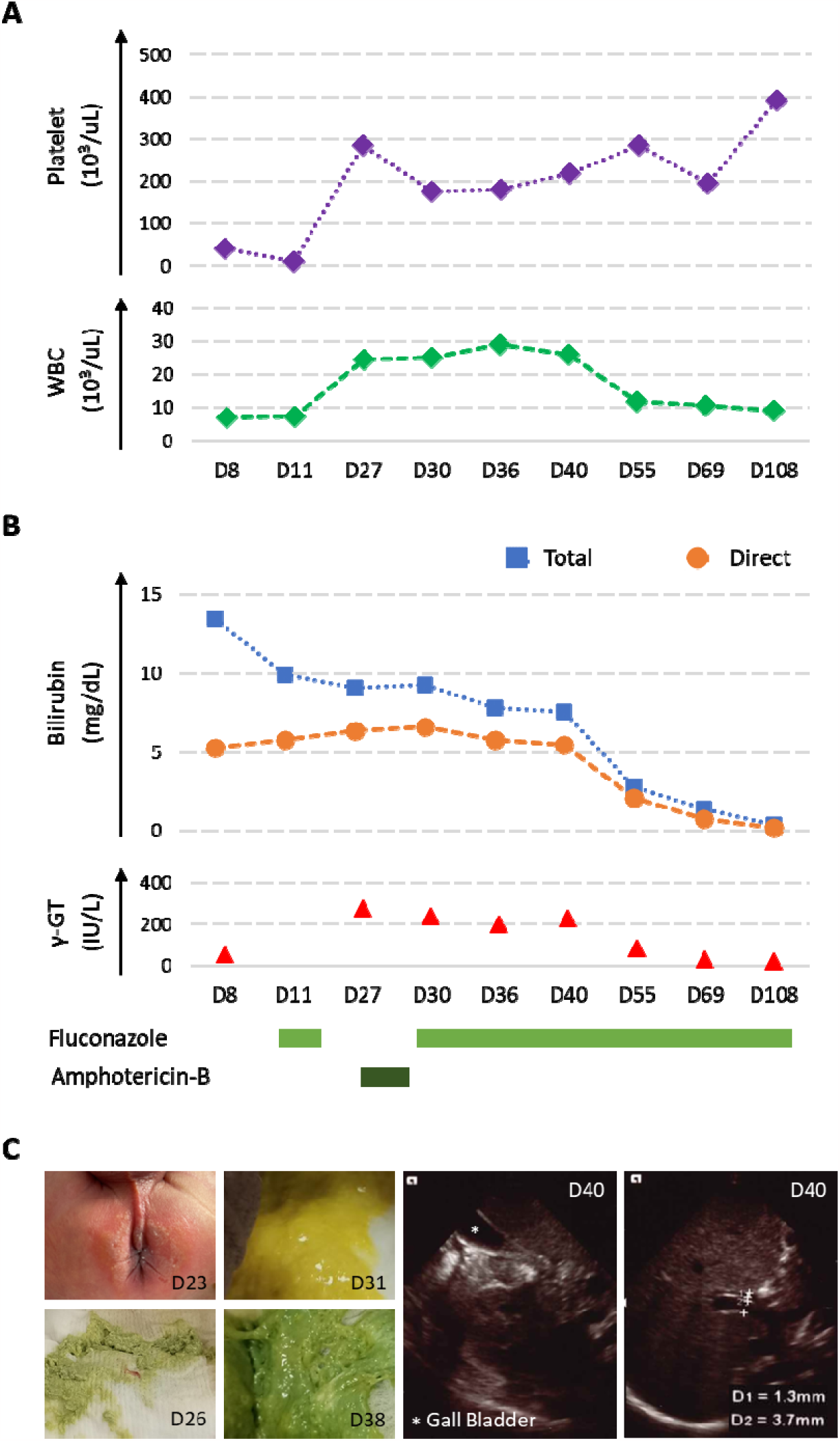
Clinical presentation of the cholangitis case. A. Profiles of platelets and white blood cell counts are distinct between early gastroenteritis stage and late cholangitis stage. B. Blood bilirubin levels and γ-GT values decrease following fluconazole treatment. C. Perianal dermatophytosis is consistent with fungal colonization in stool. A series of stool phenotypes and abdominal ultrasonographs show resolution of cholangitis.

During this period direct-type hyperbilirubinemia with high gamma-glutamyltransferase (γ-GT) values (D27, Figure 1B) was preceded by clotrimazole-responsive perianal dermatophytosis (D23, Figure 1C) and light-colored stool (D26, Figure 1C). Under suspicion of fungal cholangitis, amphotericin B was administered from D27 to D29 (Figure 1B). Without improvement on bilirubin levels, anti-fungal medication was shifted to oral fluconazole from D30 (Figure 1B). Interestingly, stool color turned to yellow on the next day (D31, Figure 1C), with presentation improving in subsequent days (D38, Figure 1C). At the same time, hyperbilirubinemia subsided gradually (Figure 1B). Abdominal echo showed a well-distended gall bladder and no bile duct obstructions (D40, Figure 1C). The patient was discharged on D41, and kept on oral fluconazole. Follow-up blood cell counts, bilirubin levels, and γ-GT values were normal (Figures 1A and 1B). Daily oral fluconazole was tapered to three times weekly for two weeks, twice weekly for two weeks, and finally once weekly for one month. The patient developed and grew normally as far as the last clinical check at five months old. Fungal cultures of blood or stool collected during the patient’s stay in the hospital all yielded negative results.

### Cholangitis associated with *Aspergillus sp*

We used fungal-ribosomal 18S PCR[4-6] to characterize suspected fungal infection from fecal DNA. Bacterial 16S V1-V9 PCR[5] served as internal control (Figure 2A). One 18S V1-V8 band was clearly amplified from the D28 fecal sample (Figure 2A), though amphotericin B was given i.v. from D27 to D29 (Figure 1B). The V1-V8 band disappeared (D31, D38, Figure 2A) after oral administration of fluconazole from D30 (Figure 1B). Products from 18S V1-V8 PCR were used to amplify the V7-V8 region for taxonomical classification through semi-nested PCR (red arrow, Figure 2A). Sequence results identified *Aspergillus sp*. by nucleotide BLAST[3] against the nr/nt database[7] (Figure 2B and Table 1).

**Table 1.**
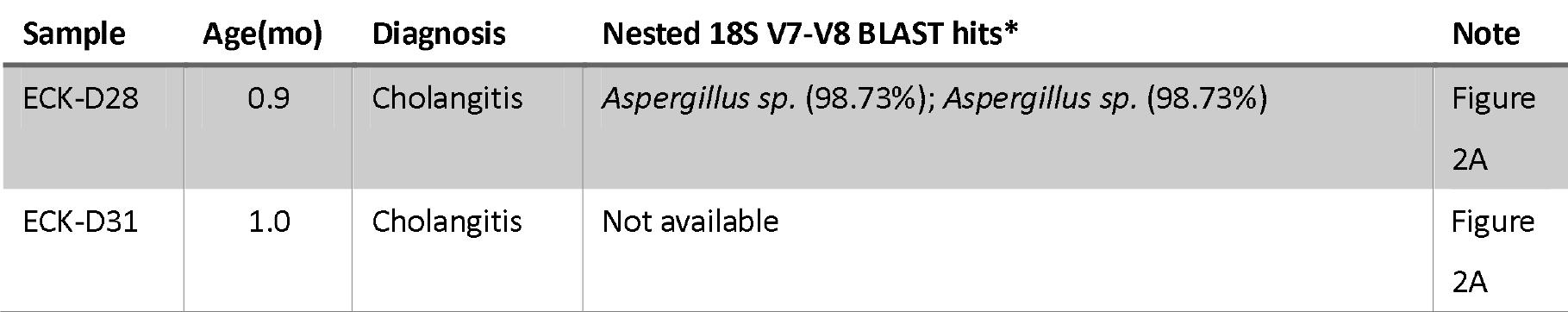
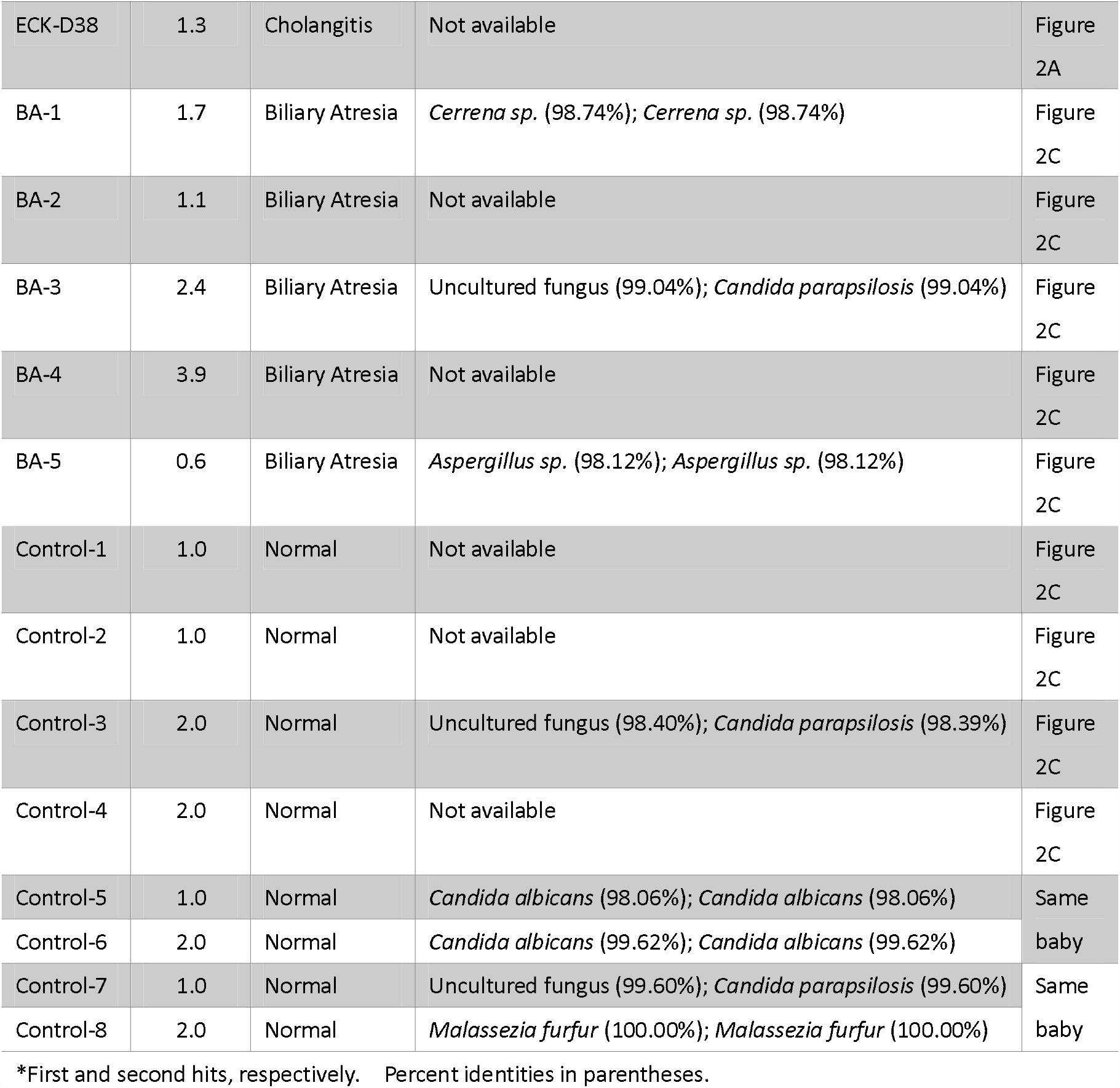
Top two hits of nested 18S V7-V8 nucleotide BLAST.

**Figure 2.**
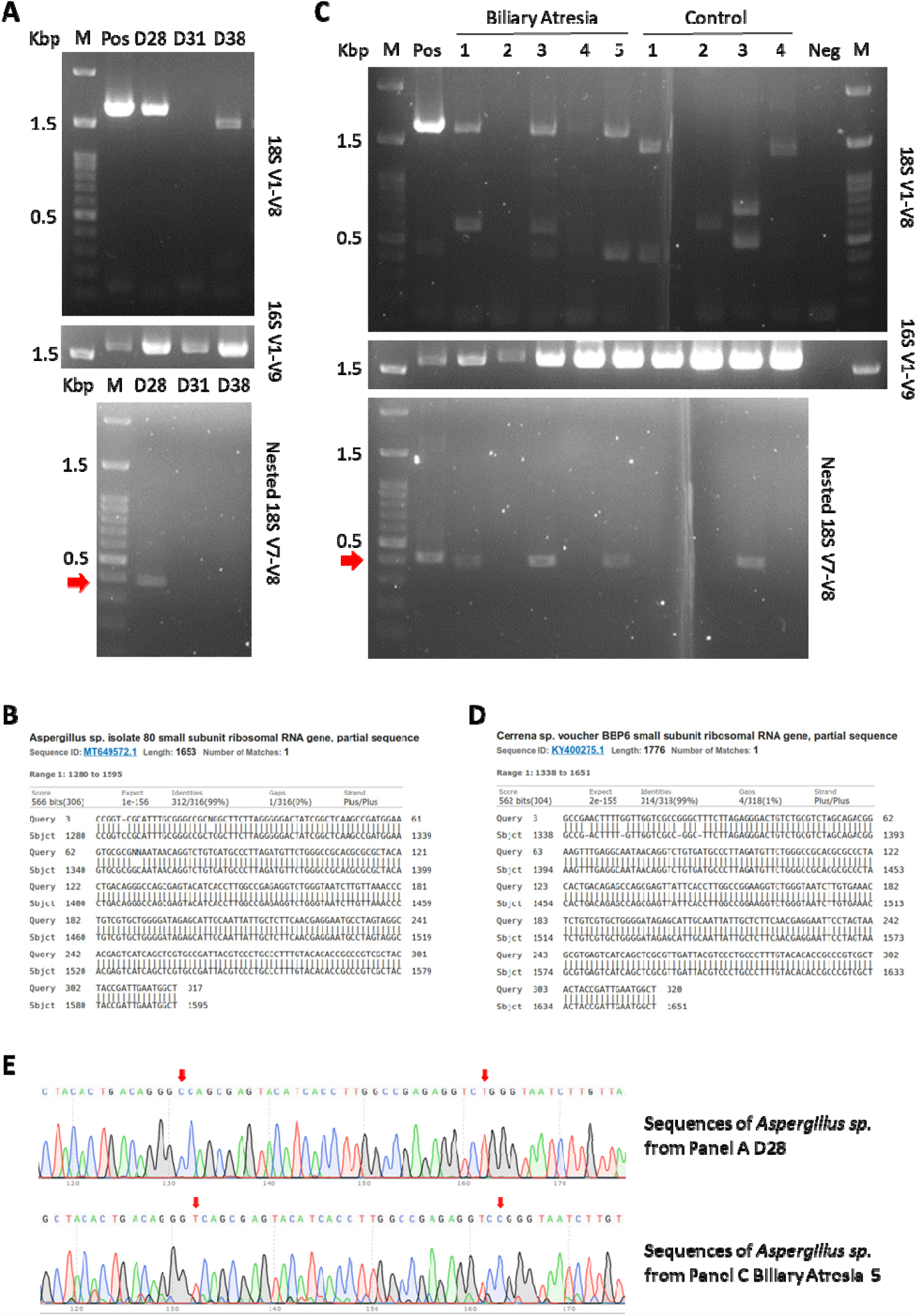
Molecular identification of fungal species with 18S ribosomal sequences. A. 18S V1-V8 and semi-nested V7-V8 (red arrow) PCRs from fecal DNA of the cholangitis case are shown. 16S V1-V9 PCR is used as internal control. B. *Aspergillus sp*. is identified in stool from the index case. C. 18S V1-V8 and semi-nested V7-V8 (red arrow) PCRs were performed against fecal DNA from BA patients and healthy controls. D. V7-V8 sequences of BA-1 match *Cerrena sp*. E. *Aspergillus sp*. are distinct between the cholangitis case and BA-5.

### Non-commensal fungal colonization in biliary atresia

*Aspergillus sp*. is not a usual fungal commensal of the gastrointestinal tract[8]. We suspect that BA, a disease also characterized by bile-duct inflammation, could be related to colonization by unusual fungi in newborn bowels. We subjected fecal DNA from five cases of BA and four healthy controls to 18S V1-V8 and nested V7-V8 PCR (Figure 2C). Four samples (BA-1, -3, -5 and control-3) yield positive results (red arrow, Figure 2C). Sequences from BA-1 are mapped to *Cerrena sp*. (Figure 2D and Table 1). *Aspergillus sp*. is also identified in BA-5 (Table 1). BA-3 and control-3 are related to Candida (Table 1). Of four additional control samples, three yield sequences from Candida and one has sequences from *Malassezia* (Table 1). Non-commensal fungi are not identified in any controls. The Fisher exact test statistic is 0.0476 (<0.05) between controls and patients who had molecular evidence of fungal colonization.

### Distinct *Aspergillus sp*. between patients

Both the cholangitis case and BA-5 are associated with bowel colonization by *Aspergillus sp*. (Table 1). We align 18S V7-V8 sequences to compare identities. We find that at least two loci have distinct T vs. C nucleotide polymorphisms (red arrow, Figure 2E). This result suggests that the two patients’ bowels were colonized by different *Aspergillus*.

## Discussion

There have been no reports of neonatal fungal cholangitis in the literature. Before the onset of hyperbilirubinemia, the index case experienced a course of gastroenteritis which responded to fluconazole but not antibiotics. Prior week-long ingestion of concentrated glucose water per os could have contributed to fungal overgrowth. Notably, the patient suffered fetal distress before emergent C/S and had a glucose level undetectable at NICU admission. Congenital fungal infection could not be ruled out.

Immune mechanisms for occurrence of cholangitis are unclear. At the gastroenteritis stage platelet counts were very low, but at the cholangitis stage platelet counts were normal (Figure 1A). Underlying pathogeneses may differ. Fungi could cause immune overreaction at the cholangitis stage but direct invasion might dominate the gastroenteritis stage. These hypotheses might be tested once culture conditions for implicated fungi, especially *Aspergillus sp*. (Figure 2B), are established.

Drawing on these findings, we suspected BA could share a fungal pathogenic mechanism. Indeed, we found colonization by non-commensal fungi in two BA patients, but not in controls. 18S analyses revealed *Cerrena sp*. in BA-1 and *Aspergillus sp*. in BA-5 (Table 1). Neither *Cerrena sp*. nor *Aspergillus sp*. are gastrointestinal commensals, though both are identifiable from human respiratory tracts[9]. These fungi might take advantage of compromised immunity in newborns to overgrow and overstimulate immature immune systems. One report describes a complicating *Aspergillus sp*. infection following Kasai portoenterostomy[10], supporting our observation of fungal overgrowth in BA patients. Though two BA cases lacked identifiable fungi, their immune systems could still have overreacted to low amounts of fungi in bowels. The index cholangitis case is an example. Although PCR did not detect fungi after fluconazole administration (D31, Figure 2A), white blood cell counts remained high for more than 10 days (Figure 1A). It is possible that trace fungal stimulants in bowels could have exponentially pathogenic effects under some circumstances.

The index case with cholangitis was successfully treated with oral fluconazole. If overgrowth and overstimulation by non-commensal fungi accounts for at least part of BA pathogenesis, early anti-fungal treatment among BA patients should be considered. Using current standard surgical interventions, two-thirds of patients still eventually require liver transplantation[2]. If BA clinical course can be significantly altered, benefits of anti-fungal treatment would clearly outweigh the small associated hepatitis risk[11].

## Data Availability

All data produced in the present study are available upon reasonable request to the authors.

## Acknowledgements

We thank Dr. Mei-Hwei Chang for professional discussion and significant support. Dr. Shiu-Feng Huang’s inputs are greatly appreciated. We thank Chih-Wei Joshua Liu (MIT Physics of Living Systems) for English editing.

## Author Contributions

S.W.H., C.J.L., and Y.H.C. conducted and optimized experiments. Y.H.N. and H.L.C. contributed samples and helped elaborate the study. H.H.L. initiated the project and proposed the hypothesis.

